# Motivation, community, and resilience in older adults’ exercise: a three-year qualitative follow-up to the BELL trial

**DOI:** 10.1101/2024.12.15.24319060

**Authors:** Neil Meigh, Alexandra Davidson, Oliver P. Thomson, Justin Keogh, Wayne Hing, Ben Schram, Sharon Mickan

**Author notes:** **Corresponding details for corresponding author**, Faculty of Health Sciences and Medicine, Bond University 14 University Drive, Robina, 4226, Queensland.

## Abstract

This qualitative study explores the long-term impact of the BELL trial’s weekly group kettlebell training on older adults’ engagement, community building, and intrinsic motivation. Three years post-trial, participants from the ‘Wednesday Friends’ group were interviewed to understand how social bonds, shared leadership, adaptability, and health benefits contributed to their sustained participation. Applying the COM-B model and the four C’s model of thematic analysis, findings reveal that social support, resilience, and self-efficacy were central to continued engagement, underscoring key motivators for older adults’ adherence to group-based exercise. These insights offer healthcare providers data-driven strategies to foster long-term engagement in community exercise programs for older adults.

## Introduction

The benefits of physical activity for older adults are widely acknowledged, with consistent evidence underscoring improvements in physical, psychological, and social well-being (Franco et al., 2015; Teixeira et al., 2012). Beyond these outcomes, physical activity fosters resilience, enhancing mental health, mood, and overall well-being, even with low-frequency interventions (Toth et al., 2024).

Despite these benefits, older adults often engage in insufficient levels of regular physical activity, especially resistance-based exercises, which are crucial for maintaining functional independence and longevity (Bauman et al., 2016; McPhee et al., 2016).

Group-based exercise programs have emerged as promising health interventions, leveraging social support, shared purpose, and community engagement to foster sustainable behavioural change (Bauman et al., 2016; Franco et al., 2015). These programs not only enhance physical outcomes but also serve as powerful tools for reducing loneliness, building self-efficacy, and promoting mental health (Bethancourt et al., 2014; Levy et al., 2020). The Ballistic Exercise of the Lower Limb (BELL) trial was designed to assess the feasibility of engaging older adults in group-based hardstyle kettlebell training, a dynamic form of resistance exercise involving a hand-held weight (Meigh, Keogh, et al., 2022). Rooted in practice-based evidence, the trial demonstrated both efficacy, in terms of health and functional outcomes, and effectiveness, as the participants demonstrated high levels of engagement and interest in continuing the program beyond the trial period (Meigh, Davidson, et al., 2022).

However, the COVID-19 pandemic caused an unexpected shift, shortening the initial in-person training phase and moving participants to an online, home-based format. It also prevented an additional 6-month training period that was planned after the trial. Following the conclusion of the trial in May 2020, some participants formed the ‘Wednesday Friends’ group, an autonomous, self-sustaining exercise group that persisted and thrived independently for three years, and continued throughout 2024.

Building on the broader conversation surrounding exercise adherence among older adults, this follow-up qualitative study situates itself within the discourse on sustainable, community-driven health interventions. Previous studies, including quantitative assessments, have evaluated attendance and physical health outcomes (Bethancourt et al., 2014; Pahor et al., 2014; Wallace et al., 1998).

However, fewer studies have explored the lived experience of older adults engaging in long-term, group-based exercise. The existing body of knowledge, often rooted in (post)positivist paradigms, offers valuable insights but leaves room for a perspective grounded in participants’ subjective experiences, social dynamics, and evolving motivations. This study employs a Critical Realist (CR) framework to provide a complementary, qualitative perspective, focusing on the reasons and mechanisms behind sustained engagement, particularly in relation to social and psychological factors (Christodoulou, 2023).

Emerging literature on group-based interventions for older adults suggests that factors such as social cohesion, shared leadership, adaptability, and resilience are crucial in fostering long-term physical activity engagement (McPhee et al., 2016; S. Smith & Johnson, 2023; Toth et al., 2024). Building on these insights, Room et al. (2017) also emphasise the importance of social support and group cohesion in sustaining exercise participation, particularly in community-based settings for older adults. Research has further shown that programs which cultivate intrinsic motivation and self-efficacy through social structures and support mechanisms are particularly effective in promoting lasting adherence to physical activity (Killingback et al., 2017). As highlighted by Cyarto et al. (2006), long-term adherence to exercise programs for older adults remains a significant challenge, and while several interventions have sought to address this, community-driven, group-based models offer promising solutions. This aligns with Keogh et al. (2014), who also noted the difficulty in sustaining exercise participation and emphasised the importance of addressing intrinsic motivation and social cohesion. This aligns with the progression observed in the Wednesday Friends group, where participants took on shared responsibility, leading to a resilient community that adapted seamlessly from structured sessions to a flexible, member-led organisation. This transition mirrors some of the findings of Wallace et al. (1998), who discussed how community-based programs can foster long-term engagement through social support and the empowerment of participants.

Self-determination theory offers a useful framework for understanding the psychological mechanisms underlying sustained engagement. Autonomously motivated behaviours, driven by a sense of competence, relatedness, and autonomy, are linked to higher long-term adherence rates (Teixeira et al., 2012). Initially supported by a structured exercise regimen, the BELL trial participants transitioned to self-directed activity, eventually becoming intrinsically motivated to maintain their fitness routines without external facilitation. This shift from guided to self-regulated behaviour exemplifies how group-based exercise can foster not only physical health benefits but also a profound shift in participant identity and self-efficacy.

As a follow-up to the BELL trial qualitative study, this study builds upon the initial insights gained regarding the physical and psychosocial benefits experienced by participants, including enhanced self-confidence, social connection, and a renewed sense of physical capability. While the BELL trial demonstrated the efficacy of kettlebell training in promoting health and functional outcomes, this study specifically focuses on understanding the continued participation and resilience of participants over time. By exploring the experience of the Wednesday Friends group, this research identifies the key drivers that enable such programs to evolve into self-sustaining communities, with a particular emphasis on the evolving role of social support, intrinsic motivation, and shared knowledge in fostering long-term healthy ageing.

## Methods

### Study Design

This study was conducted through a Critical Realist lens, positioning the research within the philosophical framework of Critical Realism (CR), which recognises the interplay between individual agency and broader social structures in shaping participants’ experiences (Fletcher, 2017). This qualitative study explores factors sustaining participation in kettlebell group training among older adults, with a focus on behaviours, self-determined motivations, and social dynamics that shape long-term engagement. Details of the initial kettlebell intervention are described in Meigh, Keogh, et al. (2022), which provides context for the exercise program participants completed.

The analysis combined the 4Cs methodology (Christodoulou, 2023) with reflexive thematic analysis (RTA) (Braun & Clarke, 2021), guided by the COM-B model (Michie et al., 2011). This integrated approach facilitated a nuanced exploration of the data, capturing the interplay between social and individual factors while structuring the findings within the COM-B framework of Capability, Opportunity, and Motivation. This CR-informed framework integrates observable behaviours and inferred mechanisms, with the COM-B model linking participants’ experiences to practical aspects of group exercise. By combining the CR perspective with RTA, we considered participant behaviours and motivations alongside social and environmental influences, ensuring a holistic, contextualised analysis of engagement in the Wednesday Friends group. Ethical approval was granted by Bond University Human Research Ethics Committee [NM03338], and written consent was obtained from participants before interviews. The study adheres to the BQQRG reporting guidelines for qualitative research (Braun & Clarke, 2024), with Supplementary File 1 providing a comprehensive overview of how the study aligns with these guidelines.

### Reflexivity

The research design and data interpretation were shaped by NM’s background in community-based group exercise interventions, prior experience leading the BELL trial, and ongoing engagement with the Wednesday Friends group since its inception. Having previously worked with participants from similar community groups and with the study participants since August 2020, NM developed interview questions that aligned with participants’ lived experiences, drawing on insights into group dynamics, social support, and motivation—key elements for long-term engagement in health programs. While this familiarity with participants’ challenges and motivations enriched the study, it also required careful attention to potential biases. Reflexivity allowed NM to critically reflect on how his prior knowledge, positions, and experience might have influenced the analysis and interpretation of the data. The ontological position of this study holds that researcher influence is inherent and beneficial, rather than something to be minimised or eliminated (Braun & Clarke, 2024). Critical reflection on NM’s role in shaping the research process and engaging with participants enabled a more nuanced, context-driven interpretation, ensuring the findings remained deeply grounded in participants’ lived experiences.

### Participants

The Wednesday Friends group represents a unique, real-world example of sustained, autonomous engagement in physical activity, offering insights into long-term adherence mechanisms in older adults. Seven participants, aged 65 to 77, included both male and female perspectives. Interviews ranged from 40 to 96 minutes in length. All active members were invited to participate to capture a comprehensive view of the group dynamics. Interviews were conducted by NM, an experienced qualitative researcher and lead investigator of the BELL trial, who has maintained contact with the group since its formation in June 2020. NM’s sustained engagement with participants in their private Facebook group, afforded a nuanced insight into individual life stories, which helped balance the benefits of this familiarity with an awareness of its influence on interpretation. The study’s purpose was explained to participants as an exploration of their ongoing engagement and motivation, with a focus on understanding social dynamics and intrinsic motivation..

### Data Generation

Semi-structured interviews were conducted in-person to accommodate participant preferences, ensuring comfort and accessibility while maintaining consistency in data generation. Questions were guided by the COM-B model, which explored how physical capability, social dynamics, and motivation contributed to long-term engagement. The interview guide (Supplementary File 2 & File 3) was designed with questions informed by the COM-B model. Interview questions underwent iterative refinement to ensure questions resonated with older adults’ perspectives on physical and social engagement in group exercise. This alignment with the COM-B model facilitated in-depth exploration of Capability, Opportunity, and Motivation factors. All interviews were audio-recorded with participants’ informed consent, and pseudonyms were assigned to ensure confidentiality. Audio-recordings of interviews were transcribed verbatim using MS Word (Microsoft Word for Microsoft 365 MSO, 2023), with identifying details removed to preserve anonymity. Intelligent verbatim transcription was used to ensure accuracy while removing non-essential fillers, reflecting participant speech patterns faithfully (Eftekhari, 2024). The dataset was sufficiently comprehensive, given the close-knit nature of the group and prolonged engagement, providing robust information power to address the study’s qualitative aims.

An inductive approach to coding (Braun & Clarke, 2021) involved the identification of patterns and topics in the data through an iterative process, where the analysis was continuously refined and adjusted as the understanding of the data deepened. Initially, data was coded in a flexible, open-ended manner, with codes generated directly from the data, guided by the study’s aim to uncover the mechanisms of participants’ engagement in the group. The coding and analysis was conducted by NM, who maintained reflexivity throughout the process, allowing for deep engagement with the data. Careful consideration was given to impartiality, ensuring that interpretations remained grounded in the participants’ perspectives. Microsoft Excel (2024) was used to support the Coding and analysis of data. R Studio was also to illustrate the connections between data (RStudio, 2024).

### Thematic Analysis Using the Four C’s Model

Thematic analysis followed Christodoulou’s Four C’s model (Christodoulou, 2023), structured in four stages: Coding, Clustering, Connecting, and Constructing (Table 1). Coded text segments from the transcribed interviews were linked to the COM-B components and practical strategies for engagement. Michie’s Behaviour Change Wheel (Michie et al., 2011) guided how COM-B components mapped to intervention functions, translating participant insights into actionable strategies. NM actively engaged with every stage of the analytic process, from coding through to the development of categories. This process was iterative, with codes being revisited and refined as new insights emerged. Through the stages of clustering and connecting, categories were linked to underlying causal mechanisms identified through retroductive reasoning. These mechanisms explain the connections between categories, shedding light on the dynamics that drive participants’ engagement in the group. The causal mechanisms are not simply thematic summaries but represent the causal dynamics that link categories, providing a deeper understanding of how different factors interact to sustain engagement over time. NM remained reflexive throughout, carefully reflecting on assumptions and biases to ensure the emerging connections were grounded in the data, faithfully representing participants’ experiences while providing meaningful theoretical insights.

**Table 1:** Stages of thematic analysis using the four C’s model for category development.

| Stage | Process and Purpose |
| --- | --- |
| Stage 1:<br>**Coding** | Transcribed interviews were reviewed and coded based on COM-B constructs and associated behaviours (e.g., capability limitations, motivational triggers). This initial coding helped identify relevant behavioural patterns and mechanisms across participant narratives. |
| Stage 2:<br>**Clustering** | Codes were grouped into categories reflecting shared practices, social dynamics, or beliefs, guided by the COM-B model. This clustering identified emerging themes related to social support, self-determined motivation, and overcoming barriers. |
| Stage 3:<br>**Connecting** | Social network analysis was employed to visualise the connections between categories. Categories were linked if one influenced, enabled, or conditioned another, illustrating the interdependencies within the group's experience. |
| Stage 4:<br>**Constructing** | Overarching themes were developed by redescription of category connections. A hierarchical structure was created to highlight core themes that significantly impact sustained engagement and group cohesion. |

## Analysis

The clustering of codes produced ten categories, which were integrated into a unified one-mode network capturing the participants’ experiences in the weekly group training. Together, these categories offer a comprehensive understanding of participants’ experiences within the kettlebell training group. They also form the basis for developing broader themes that capture the key dynamics shaping participants’ engagement.

### Stage 1 - Initial / In vivo Coding

A total of 364 unique codes were generated from text fragments across the transcribed interviews: Sophia (71), Quinn (64), Megan (53), Kimberly (47), Julia (49), Felicity (51), Derek (29). The detailed clustering of codes into categories is presented in Supplementary File 4.

#### Sources of Behaviour & Intervention Functions

The three domains of capability, opportunity, and motivation were nearly evenly represented across the codes identified in the transcribed interviews, suggesting that each domain played an essential role in participant engagement and behavioural persistence within the group. Further analysis of the components of the COM-B revealed notable differences within these primary domains – *social opportunity* was a dominant factor (103 codes, 28%), while *physical opportunity* accounted for considerably fewer codes (21 codes, 6%). Eighteen of the possible 21 COM-B component and intervention function combinations were present in the coded data, with five combinations accounting for the majority (57.7%) of the codes. Representative participant data are provided in Supplementary File 5.

The most prominent intervention functions, derived from the behavioural domains of the COM-B model and identified in the data, are summarised in Table 2. These intervention functions align with the behaviours that most strongly contributed to sustained engagement in the Wednesday Friends group. This analysis provides valuable insights into the core factors that support long-term participation, offering guidance for future interventions aimed at promoting similar behavioural changes in community-based programs for older adults.

**Table 2:** Most dominant intervention functions derived from COM-B behavioural domains.

| Intervention Function | Description |
| --- | --- |
| Enablement + Environmental Restructuring (*social opportunity*) | Emphasising the role of social cohesion and mutual support in facilitating sustained engagement, with a focus on shared responsibilities, inclusivity, and structured group dynamics that make participation accessible and enjoyable (16%). |
| Education (*psychological capability*) | Reflecting participants' appreciation for structured education on exercise techniques, fostering a nuanced understanding of how physical activity benefits health, enhances self-efficacy, and supports positive ageing identity (13%). |
| Training (*physical capability*) | Focusing on structured, adaptive training methods that empowered participants to build essential physical skills, such as strength, balance, and resilience, needed for effective participation in group exercises (10%). |
| Education (*reflexive motivation*) | Reinforcing motivation through knowledge-sharing and participant-driven encouragement, enhancing self-efficacy and personal commitment to sustained participation (7%). |

### Stage 2 – Clustering

Clustering of codes resulted in ten unified categories, summarised in Table 3. These combine the observed (what was said by participants and read in the transcribed interviews) and unobserved (shared meaning between codes).

**Table 3:** Summary of unified categories explaining sustained participant engagement.

| Code | Category | Explanation |
| --- | --- | --- |
| C1 | Social cohesion and support | Encompasses topics around social bonding, mutual encouragement, shared experiences, and friendship – Codes that highlight social dynamics, the importance of group bonds, and the self-determined motivation derived from being part of a collective are included here. |
| C2 | Mental and emotional well-being | Focuses on psychological benefits, emotional support, and the impact of exercise on mental health – Codes reflecting changes in emotional state, the psychological uplift from exercise, and mental resilience are grouped under this category. |
| C3 | Leadership and group dynamics | Addresses the influence of leadership, the role of key individuals, and how group roles are negotiated – Includes codes related to leadership identification, dependency, and the collaborative or self-directed nature of the group. |
| C4 | Motivation and commitment | Captures intrinsic and extrinsic motivations, personal drive, and adherence to routines – Codes detailing the reasons for continued participation, dedication to training, and factors influencing self-determined motivation belong here. |
| C5 | Physical health and capability | Relates to physical fitness, health management, improvements in capability, and dealing with physical limitations – Codes covering physical outcomes, overcoming health challenges, and the tangible benefits of physical activity are placed in this category. |
| C6 | Practical considerations and flexibility | Includes logistical aspects, adaptability in training, and accessibility – This category gathers codes about logistical challenges, accommodating diverse physical abilities, and the flexible nature of participation. |
| C7 | Skill development and knowledge sharing | Encompasses the importance of proper technique, learning, and transferring knowledge – Codes discussing technical skill acquisition, confidence in teaching, and sharing resources are organised here. |
| C8 | Behavioural change and maintenance | Focuses on sustained behaviour change, self-efficacy, and the impact of regular exercise – Includes codes reflecting on changes in daily habits, persistence in maintaining routines, and long-term behavioural shifts. |
| C9 | Identity and self-perception | Covers how participation affects self-image, identity, and perception of ageing – Codes that highlight personal growth, changes in self-perception, and the evolving sense of self are categorised here. |
| C10 | Challenges and barriers | Examines the obstacles faced, including health barriers, social challenges, and group attrition – Codes addressing dropout rates, individual struggles, and limitations encountered during the training process are included. |

### Stage 3 – Connecting

A one-mode network (Supplementary file 6) was developed to examine the relationships between the ten unified categories, highlighting their relational and emergent properties within the context of the Wednesday Friends group. The relational map (Figure 1) visualises these connections, enabling an analysis of which categories demonstrate the highest salience of connectivity and are therefore central to participants’ experiences. Categories with significant relational influence, such as *motivation and commitment* (C4) and *challenges and barriers* (C10), emerged as having the greatest salience, given their direct and enabling effects on other aspects of group engagement.

**Figure 1:**
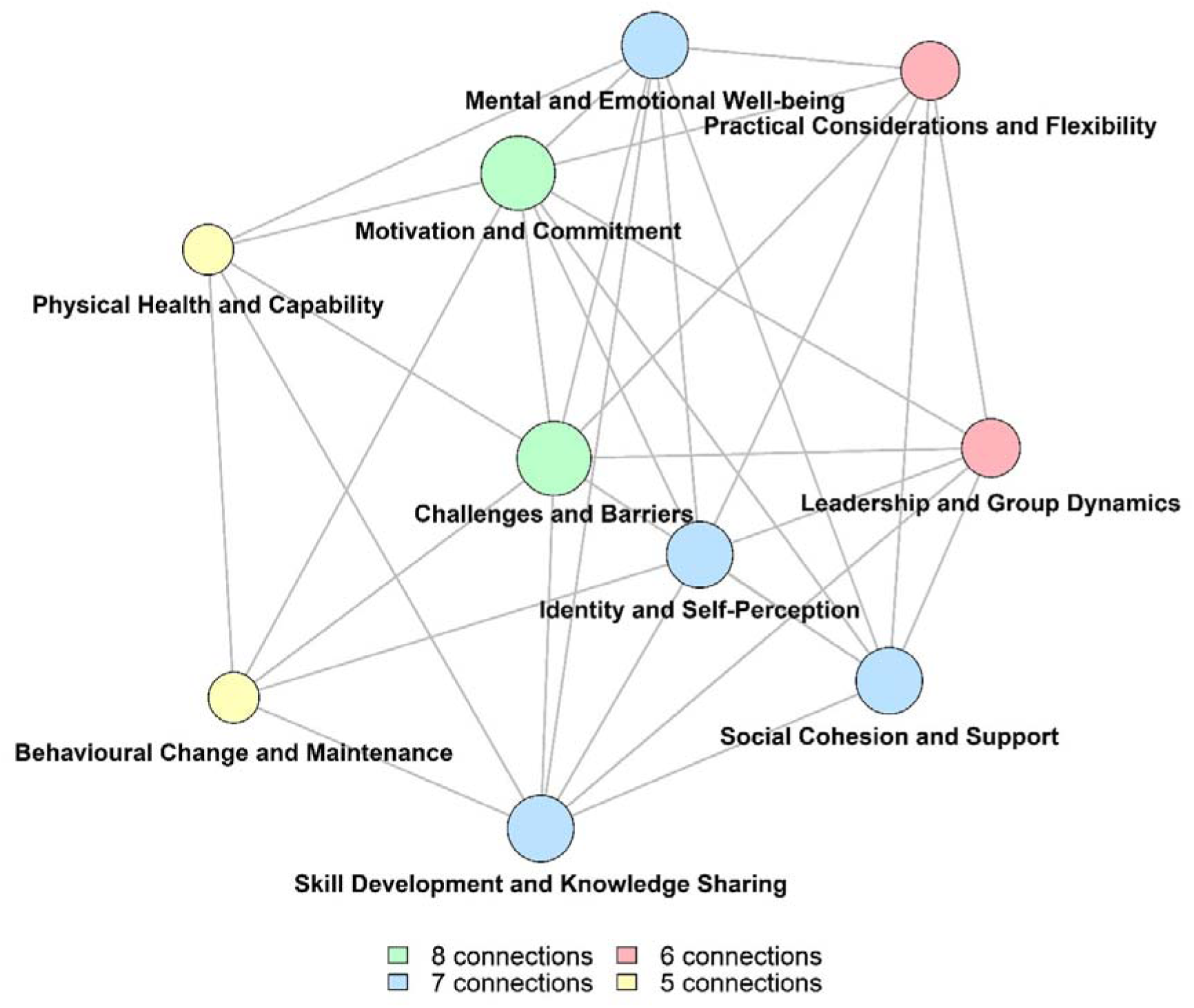
Relational map depicting connections between categories.

Key insights from these connections highlight that *motivation and commitment*, along with addressing *challenges and barriers*, are pivotal in sustaining engagement, as they significantly influence how participants perceive and interact with the group. Similarly, *social cohesion and support*, *mental and emotional well-being*, and *skill development and knowledge sharing* are strongly associated with personal engagement and satisfaction within the group. Supplementary File 7 provides a detailed explanation of the category connections and the rationale behind their relationships, illustrating how these elements interact within the context of the Wednesday Friends group.

This network analysis offers a comprehensive perspective on the interconnections that shape the group’s experience, identifying areas that could serve as focal points for designing or enhancing group-based interventions. The following sections will expand on these findings, offering deeper insights into how these relationships support sustained engagement among the Wednesday Friends group.

### Stage 4 – Constructing

Through the analysis of category connections within the one-mode network, a structured hierarchy emerged, guiding the construction of four core themes. The connections between categories reveal how each aspect of the group exercise experience interacts with others, with some categories emerging as central to sustaining engagement and fostering behavioural change. These core connections provide a foundation to understand the mechanisms that drive group dynamics and maintain personal commitment.

Based on connectivity:

(a) C4 (motivation and commitment) and C10 (challenges and barriers) emerged as the most interconnected categories, each attracting eight connections, indicating their foundational roles in sustaining participation despite obstacles.
(b) C1 (social cohesion and support), C2 (mental and emotional well-being), C7 (skill development and knowledge sharing), and C9 (identity and self-perception) formed the next level of influence, each exhibiting seven connections, reflecting their roles in fostering a supportive group environment and enhancing individual engagement through social and psychological pathways.
(c) C3 (leadership and group dynamics) and C6 (practical considerations and flexibility) demonstrated six connections, highlighting their crucial, albeit slightly less central, roles in providing structural and practical foundations for the group’s sustained functioning.
(d) C5 (physical health and capability) and C8 (behavioural change and maintenance) attracted five connections, indicating their integral contributions to maintaining health and reinforcing long-term behavioural change

These relationships led to the construction of four major themes, each reflecting a distinct yet interconnected aspect of the group exercise experience:

**THEME 1** (connectivity (a)): **motivation and resilience as central to sustaining participation**

The strong link between *motivation and commitment* (C4) and *challenges and carriers* (C10) underscores the critical role of resilience in sustaining long-term engagement. Motivation, both personal and collective, provides the driving force that helps participants navigate physical, emotional, and logistical obstacles. Mutual encouragement strengthens members’ personal commitment to both the group and their own well-being, fostering a collective resilience that addresses setbacks together. Derek’s statement, “If I don’t do it, I just feel absolute crap” (CD6), powerfully conveys the emotional and physical importance of participation, showing how deeply self-determined motivation is intertwined with both personal health and the group’s collective commitment to supporting each other through challenges.

**THEME 2** (connectivity (b)): **social and psychological interactions as drivers of engagement**

The connections between *social cohesion* (C1), *mental and emotional well-being* (C2), *skill development and knowledge sharing* (C7), and *identity and self-perception* (C9) highlight how social and psychological interactions underpin ongoing engagement and identity within the group. These connections reflect a supportive social environment that not only strengthens individual self-confidence but also builds a shared identity, deepening personal commitment to group exercise. As members grow in competence through shared learning and mutual support, they also gain a stronger sense of belonging. Sophia’s words, *“I’d be lost without it”* (CS19), reflect how group cohesion offers an anchor that extends beyond physical training, reinforcing the group’s role as a source of emotional stability, encouragement, and shared purpose.

**THEME 3** (connectivity (c)): **effective leadership and practical adaptability as foundations for group success**

The connection between *leadership and group dynamics* (C3) and *practical considerations and flexibility* (C6) shows how adaptable leadership and flexible structures provide a stable foundation for ongoing participation. Leaders who balance guidance with inclusivity foster an adaptable environment where all members feel empowered to contribute. This flexibility ensures that the program remains accessible, reducing barriers and dropout rates. Sophia’s reflection, “Sometimes you have to fake it ’til you make it” (CS17), captures both the emotional and practical challenges that group members face, underscoring the importance of adaptable leadership in maintaining group cohesion, especially as members support one another in overcoming personal and logistical hurdles.

**THEME 4** (connectivity (d)): **health improvements as a reinforcement for long-term behavioural change**

The connection between *physical health and capability* (C5) and *behavioural change and maintenance* (C8) illustrates how health gains act as powerful reinforcements for sustaining new habits. Tangible improvements in strength, endurance, and overall physical health not only validate participants’ efforts but also motivate them to maintain their personal commitment to the group. This interplay between health benefits and behavioural consistency is crucial, as health improvements offer both a reward and a reminder of the value of participation. Sophia’s comment, “I just feel that I can do anything still, I don’t feel any different to when I was 35” (CS15), embodies the sense of empowerment and positive self-perception that comes from consistent exercise, showing how health improvements foster both physical and mental resilience.

These themes capture the key social, psychological, practical, and health factors that keep the Wednesday Friends group engaged and committed. Together, they offer a clear framework for understanding what drives long-term participation, showing how group dynamics and personal motivations come together to create a lasting, supportive community.

In the final stage of analysis, we employed retroductive reasoning to uncover the key mechanisms driving the Wednesday Friends group’s experience (Figure 2). Using a CR approach, we went beyond surface-level details to identify four core mechanisms that illustrate how various aspects of participants’ experiences are interrelated and collectively shape their engagement in the weekly kettlebell training sessions. These mechanisms are not merely descriptive but reflect the deeper relational connections between categories, offering a causal explanation of the factors that sustain long-term involvement.

**Figure 2:**
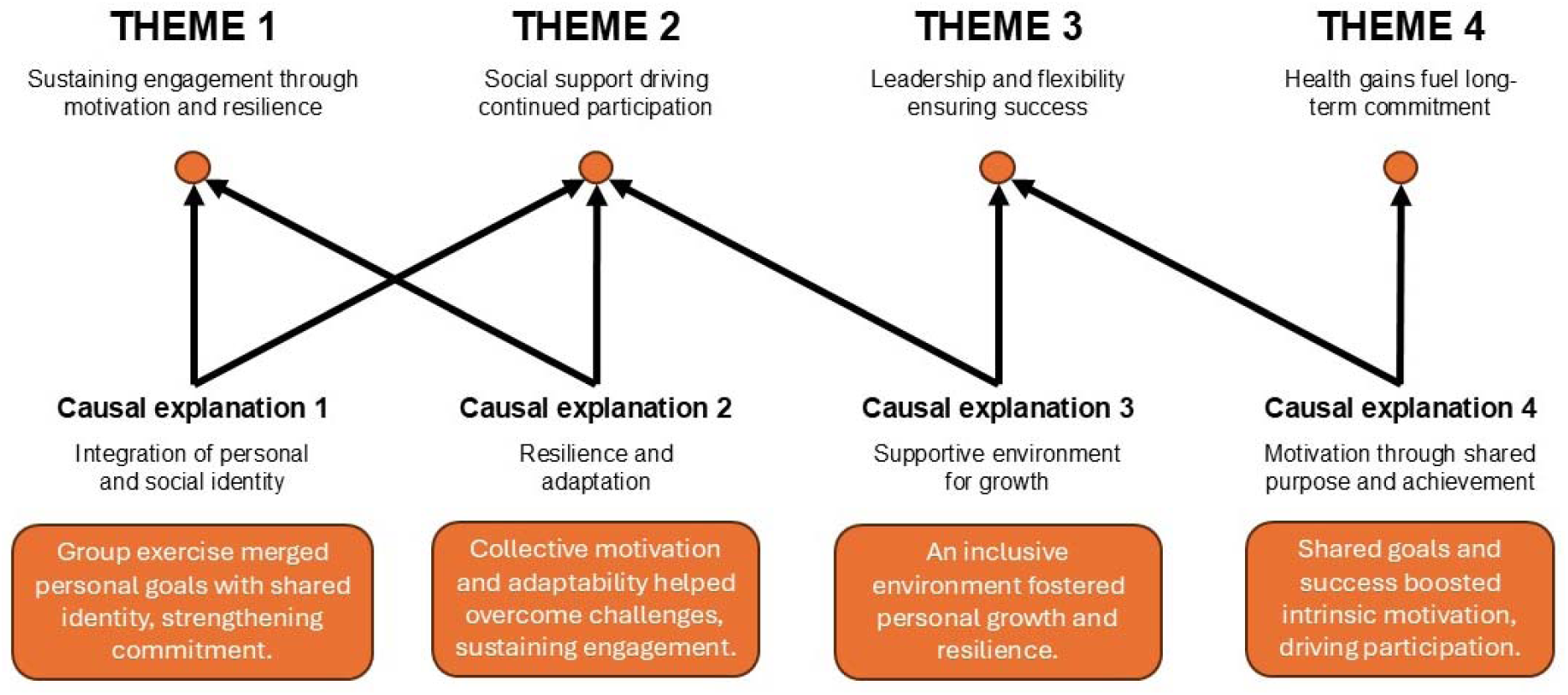
Retroductively inferred causal explanations of themes.

#### Causal explanation 1: the integration of personal and social identity through group exercise (Themes 1 & 2)

During the BELL trial, group exercise sessions enabled participants to merge personal identity (individual health goals, challenges, and experiences) with a collective social identity (belonging to the group). The cohesive support network, encouragement, and shared purpose led to a heightened sense of personal and collective identity. This merging is critical because it transformed exercise from an individual, often isolated, activity into a socially-driven experience, reinforcing personal commitment and enhancing self-perception. Many participants described feeling a strong sense of belonging, motivation from the group, and a transformation in how they viewed themselves as they continued their journey. Statement like “It’s just a good outlet. As you know, I always trained anyway, and now it got me back into [feeling like] if I don’t train I feel guilty” underscore the integration of this social influence into a newly developed self-identity.

#### Causal explanation 2: resilience and adaptation in the face of challenges (Themes 1 & 2)

Resilience and adaptability became crucial mechanisms, allowing participants to continue despite physical, emotional, or logistical challenges. Group exercise fostered a form of shared resilience, where individual struggles were mitigated by collective support. Adaptability, particularly the flexibility of exercise routines and schedules, enabled members to find ways to remain engaged despite life’s obstacles. This collective resilience, supported by the group’s social fabric, was vital in sustaining participation over the long term.

Participants’ accounts often highlighted the social and emotional support received during difficult times. For example, Derek shared, “Everyone motivates everyone, that’s the thing” reflecting the mutual motivation that exists within the group. This peer encouragement, rooted in a shared sense of commitment, was essential in helping individuals adapt to challenges and maintain their engagement, even in the face of adversity.

#### Causal explanation 3: the power of a supportive environment for personal growth (Themes 2 & 3)

The supportive environment created by the group fostered a sense of safety and encouraged personal growth and development. Within this context, individuals were able to take on challenges they might have otherwise avoided and develop new skills, contributing to both physical and emotional well-being. This sense of safety and encouragement was not accidental but emerged from the intentional practices of inclusivity, non-judgemental guidance, and shared learning fostered by the group dynamics. For example, one participant recalled, “We did exercises the other day when Sophia turned around and said ‘I’ve got a circuit for us’, and it was a lot of leg work” emphasising how group guidance and structured exercises helped participants feel both challenged and supported. This environment allowed for individual growth within a collective space, enhancing both physical health and mental resilience.

#### Causal explanation 4: motivation driven by shared purpose and collective achievement (Themes 3 & 4)

The shared purpose of achieving health goals, coupled with visible group achievements, acted as a motivational catalyst for continued engagement. The ability to adapt and modify exercise routines without sacrificing the sense of progress was essential. This flexible approach allowed participants to experience personal achievement while contributing to the collective success of the group. Over time, this sense of shared purpose and visible achievements helped reinforce positive identities and elevated the perceived value of participation. Examples such as Sophia’s comment, “We would never have gelled if it wasn’t for kettlebells” highlight the importance of the shared activity in fostering group cohesion. This bond, built through mutual goals and collective achievements, motivated participants to persist and adapt, reinforcing both personal and group identities. By recognising these underlying causal mechanisms, we can appreciate how these themes are not merely isolated concepts but interconnected phenomena that, together, explain the sustained engagement and success of the BELL trial’s group exercise program.

As the data suggests, the group’s experience evolved from structured exercise into a small but vibrant community. Over time, members displayed an unwavering personal commitment not only to their own physical health but to each other’s well-being, transforming exercise into a collective journey of resilience and identity building.

## Discussion

This study highlights the essential role of social support, motivation, and shared responsibility in sustaining long-term participation in group-based exercise programs for older adults. The Wednesday Friends group exemplifies how these elements—motivation, social cohesion, and resilience—interact to foster continued engagement in physical activity over time. The findings emphasise the complex and interconnected nature of the factors that sustain participation, demonstrating that successful group-based exercise programs are those that not only promote physical health but also nurture the psychological and social aspects of well-being.

The analysis identified key themes related to motivation, social dynamics, and practical considerations, all of which contribute to maintaining long-term engagement. As participants in the Wednesday Friends group transitioned from a structured exercise regimen to a more flexible, self-directed model, they demonstrated increasing intrinsic motivation. This shift is consistent with Self-Determination Theory (Ryan & Deci, 2000), which posits that behaviours driven by intrinsic motivation are more likely to result in sustained engagement. The group’s ability to adapt and self-regulate— fostering a sense of autonomy—was pivotal in maintaining their commitment to the program, underscoring the importance of building environments that support these psychological drivers of engagement (Bonanno, 2004)

These findings contribute to the growing body of literature on long-term exercise adherence by offering insights into the shift from extrinsic to intrinsic motivation among older adults. In line with Jones and Musgrave’s (2024) recent recommendations for successful group fitness programs, this study found that social support and the creation of a cohesive, supportive environment were key to sustaining engagement in group exercise among older adults. The Wednesday Friends group’s experience reinforces the importance of fostering a strong sense of community and belonging, which in turn facilitates continued participation. This aligns with previous studies suggesting that social support structures—coupled with the empowerment of participants—are integral to long-term engagement in physical activity (Cohen & Wills, 1985; Cyarto et al., 2006; Di Lorito et al., 2021; Farrance et al., 2016; Room et al., 2017; G. L. Smith et al., 2017).

Self-determination theory further illuminates how group cohesion, autonomy, and competence contribute to intrinsic motivation, supporting sustained adherence. The transition from externally driven participation to self-motivated engagement within the Wednesday Friends group illustrates the critical role of a supportive social environment in developing self-efficacy and personal commitment. This process of transformation highlights how group-based exercise programs can lead to profound shifts in participants’ self-perception and sense of ownership over their health, which are crucial for long-term success (Ryan & Deci, 2000). As early successes in physical improvements catalyse intrinsic motivation, as noted by Cross et al. (2023), the social context of the group further reinforces this self-driven commitment, promoting continued physical activity engagement (Rodrigues et al., 2023; Tao et al., 2024).

The COM-B model offers an actionable framework for healthcare providers aiming to foster sustainable engagement in group-based exercise programs for older adults. The following intervention functions, derived from our data, offer actionable pathways to design impactful, community-based interventions:

1. Enablement + environmental restructuring (*social opportunity*) – A supportive social structure and a conducive physical environment are essential for sustaining long-term engagement. In the Wednesday Friends group, community-building, inclusive locations, and peer interaction were critical for reinforcing social support and consistent participation. Healthcare providers should prioritise creating accessible, welcoming environments with regular schedules, designed to foster both social engagement and convenience. Research supports that these environments can significantly enhance attendance and participation, reduce barriers to engagement, and build a collective sense of purpose (Weselman et al., 2022).
2. Education (*psychological capability*) – Participants’ appreciation for structured education on exercise techniques highlights the value of educational initiatives in enhancing self-efficacy. Programs should incorporate workshops on safe exercise techniques and goal-setting sessions to improve participants’ confidence and capability. Studies by Young et al. (2024) and Meredith et al. (2023) support the role of education in building the skills necessary for long-term, autonomous engagement in physical activity.
3. Training (*physical capability*) – Structured, progressive training programs that focus on skill-building are key to sustaining engagement. By offering exercises that increase physical competence, healthcare providers can instil a sense of achievement and boost confidence. Research by Burton et al. (2022) confirms that skill-building not only improves physical function but also promotes long-term adherence by fostering a sense of accomplishment and mastery.
4. Education (*reflexive motivation*) – Knowledge-sharing and peer-driven encouragement enhanced reflective motivation within the group. These interactions reinforced participants’ personal achievements and fostered intrinsic motivation. Incorporating opportunities for group knowledge-sharing can build self-determined motivation, supporting long-term participation. As noted by Mappanasingam et al. (2024), recognition and shared experiences within a group strengthen motivation and enhance engagement.

While the success of the Wednesday Friends group provides valuable insights, there are several factors that should be considered when generalising these findings. The study focused on a specific cohort of older adults participating in a community-based kettlebell exercise program, which may limit the applicability of the results to other populations or exercise modalities. The sample was predominantly homogeneous, consisting entirely of Caucasian participants, and future research could explore how these findings apply to more culturally diverse groups or those with varying health conditions. Nevertheless, the intervention functions identified in this study—social opportunity, psychological capability, physical capability, and reflexive motivation—offer exciting potential for application across a broad spectrum of current and emerging exercise programs for older adults.

These functions could be integrated into a range of community-based programs, health and fitness professional-led programs, and workplace wellness initiatives, creating a framework for fostering sustained engagement and promoting healthy ageing in diverse settings.

## Conclusion

This study reinforces the critical role of social support, shared responsibility, and intrinsic motivation in fostering long-term engagement in group-based exercise programs for older adults. The findings highlight the importance of creating flexible, supportive environments that promote autonomy, competence, and social connectedness. These factors not only enhance physical health but also contribute to the psychological well-being and resilience of older adults, making it more likely that they will continue to engage in exercise over time. Policymakers and healthcare providers should consider these findings when designing community-based exercise programs, ensuring that they foster an environment of social cohesion and mutual support that encourages sustainable participation.

## Data Availability

All data produced in the present work are contained in the manuscript

## Acknowledgements

The authors sincerely thank the participants who generously shared their experiences of the Wednesday Friends group kettlebell training. Thanks also to Carly Hudson for assisting in the creation of the network map.

## Author’s contributions

CRediT author statement - **Neil Meigh**: Conceptualisation, Data curation, Formal analysis, Investigation, Methodology, Project administration, Software, Resources, Validation, Writing – original draft, – review & editing. **Alexandra Davidson**: Conceptualisation, Methodology, Writing – review & editing. **Oliver P. Thomson**: Conceptualisation, Methodology, Writing – review & editing. **Justin Keogh**: Methodology, Writing – review & editing. **Wayne Hing**: Methodology, Writing – review & editing. **Ben Schram**: Methodology, Writing – review & editing. **Sharon Mickan**: Conceptualisation, Methodology, Supervision, Writing – review & editing.

## Availability of data and materials

All data generated or analysed during this study are included in this published article. Bond University Human Research Ethics Committee has not authorised the public release of participants’ transcribed interviews.

## Declarations

### Ethics approval and consent to participate

All research activities were conducted in accordance with relevant guidelines and regulations, in accordance with the Declaration of Helsinki. The study was approved by the Bond University Human Research Ethics Committee (BUHREC; NM03338). Informed consent was obtained by the lead investigator from all participants.

### Consent for publication

Not applicable.

