## Supplementary File 7 for "Motivation, community, and resilience in older adults’ exercise: a three-year qualitative follow-up to the BELL trial" (from another version of this preprint: version 1, 2024-12-16)

##### Supplementary File 7: Relationality between categories

**C1: Social Cohesion and Support**

- **C1 - C2: Social cohesion reinforces mental and emotional well-being through the supportive network.**
  In a cohesive group, the sense of belonging and mutual support contributes directly to improved emotional well-being. For example, Derek mentioned how being part of a group that "pushed each other" made him feel supported, which positively influenced his mood and reduced feelings of isolation. This highlights how the group's social dynamics help alleviate stress and enhance overall mental resilience.
- **C1 - C3: Leadership and group dynamics emerge as a key part of social cohesion, fostering a sense of belonging.**
  Effective leadership plays a significant role in creating a supportive social environment. Leaders like Sophia, who subtly guided the group without overt dominance, allowed members to feel equally valued, thereby fostering cohesion. This type of leadership helps to establish norms of participation and mutual respect that are crucial for a tight-knit community.
- **C1 - C4: Motivation is enhanced by the supportive group, as encouragement and social bonds drive commitment.**
  Derek’s internal motivation to attend sessions was closely tied to the group’s supportive structure. He expressed that seeing others commit to the training reinforced his own commitment. The social context provided both accountability and encouragement, key elements for maintaining motivation over time.
- **C1 - C6: Practical aspects like scheduling and location flexibility are integral to maintaining social cohesion.**
  Flexibility within the group’s schedule and training environment, such as adapting to outdoor locations when necessary, helped maintain social cohesion by accommodating members’ diverse needs. For example, the group's willingness to meet at familiar locations even under adverse conditions highlights the adaptability that preserves social ties.
- **C1 - C7: Social cohesion encourages knowledge-sharing and skill development within the group.**
  The camaraderie fostered by social cohesion leads to a culture of collective learning. Felicity often shared her training insights and progress with other group members, leading to a reciprocal exchange of tips and techniques. This open sharing not only develops individual skills but also strengthens the group’s knowledge base.
- **C1 - C9: A strong sense of identity and self-perception is fostered through positive group interactions.**
  Members often described how being part of the group positively influenced their self-image. Quinn, who initially struggled with self-confidence, began to see himself as a competent and valued member due to the group's encouragement, illustrating how cohesive social interactions can shape self-perception.
- **C1 - C10: Challenges and barriers are easier to overcome in a cohesive group, providing resilience.**
  Group cohesion acted as a buffer against obstacles. When Felicity faced health issues, the group’s consistent support encouraged her to return and continue training. This example shows that the group's collective encouragement can help members navigate setbacks, making challenges more manageable.

**C2: Mental and Emotional Well-being**

- **C2 - C1: Mental well-being benefits from the supportive environment provided by social connections.**
  A participant like Derek, who dealt with emotional challenges, found solace in the consistent and positive environment offered by the group. The sense of camaraderie reduced feelings of isolation and anxiety, underlining how social support is a critical factor in maintaining emotional stability.
- **C2 - C4: Emotional well-being fuels motivation, helping members remain committed to group activities.**
  Positive emotional experiences during sessions—such as the joy of shared accomplishments—served as strong motivational factors. Sophia’s excitement about the group’s progress boosted her commitment, showing how emotional satisfaction can reinforce long-term engagement.
- **C2 - C5: Physical health improvements positively impact emotional well-being, enhancing mental resilience.**
  Members like Kimberly reported that as their physical health improved, they noticed enhanced mood and mental clarity. This relationship between physical health and emotional stability suggests that physical gains contribute significantly to overall well-being.
- **C2 - C6: Flexibility and practical considerations play a role in reducing mental stress and maintaining well-being.**
  Reducing logistical pressures through adaptable scheduling mitigates stress. Participants appreciated the group's ability to adjust for individual circumstances, like accommodating different paces and preferences, thus creating a relaxed atmosphere conducive to well-being.
- **C2 - C7: Mental well-being is connected to skill development, as acquiring new skills can enhance self-efficacy.**
  Learning new skills provided members with a sense of competence and accomplishment. For instance, Felicity’s pride in mastering kettlebell techniques increased her confidence and reduced anxiety, demonstrating how skill acquisition contributes to mental well-being.
- **C2 - C9: Improved mental and emotional well-being is tied to enhanced self-perception and personal identity.**
  Feeling mentally and emotionally secure allowed participants to perceive themselves more positively. Quinn, who initially saw himself as physically limited, reported feeling more capable and less anxious as his emotional well-being improved through consistent group support.
- **C2 - C10: Addressing mental barriers is crucial for maintaining emotional health, aiding in overcoming challenges.**
  Challenges like self-doubt and anxiety were openly discussed within the group, helping individuals like Megan confront and overcome their mental barriers. The group’s approach to openly addressing these challenges reinforced emotional health and facilitated personal growth.

**C3: Leadership and Group Dynamics**

- **C3 - C1: Effective leadership shapes group cohesion and supports social bonds.**
  Leaders played a pivotal role in nurturing a cohesive environment. For example, Sophia’s balanced guidance, which encouraged autonomy while providing support, cultivated trust and connectedness among group members, reinforcing the group’s unity.
- **C3 - C4: Leadership styles influence motivation, inspiring members to stay engaged.**
  The leadership approach had a direct impact on motivation. Sophia’s method of acknowledging individual achievements and providing constructive feedback motivated members to persist. This illustrates how a leadership style that validates effort can inspire sustained participation.
- **C3 - C6: Practical group management requires effective leadership, ensuring activities run smoothly.**
  Effective leaders manage logistical challenges by organising sessions and adapting to unforeseen circumstances. Quinn’s facilitation of schedule changes during weather disruptions showed how practical leadership supports the group’s overall function and participation.
- **C3 - C7: Leaders facilitate skill-sharing, making leadership integral to knowledge development.**
  The leader’s role in encouraging skill exchange was crucial. Kimberly often led impromptu mini-sessions to demonstrate techniques, emphasising the importance of leadership in fostering a learning culture that benefits the entire group.
- **C3 - C9: Leadership helps shape group identity and fosters positive self-perception among members.**
  Strong leadership helped members see themselves as integral parts of the group. Derek mentioned how the leader’s validation made him feel like a valued participant, enhancing his sense of identity and belonging within the group.
- **C3 - C10: Good leadership can navigate and mitigate challenges faced by the group.**
  Leaders often acted as problem-solvers. When logistical or interpersonal challenges arose, like conflicting schedules, strong leadership ensured that solutions were found quickly, minimising disruption and maintaining group morale.

**C4: Motivation and Commitment**

- **C4 - C1: Social support reinforces motivation, driving ongoing commitment.**
  Encouragement from the group provided a sense of accountability, with members like Megan reporting that the supportive environment was crucial in maintaining her motivation to attend sessions even during low periods.
- **C4 - C2: Mental resilience contributes to motivation, helping maintain commitment.**
  Stable mental health underpins consistent motivation. Kimberly noted that the improvement in her emotional stability gave her the energy and mental capacity to stay committed to the training, even when faced with physical fatigue.
- **C4 - C3: Leadership plays a pivotal role in sustaining motivation and adherence.**
  The presence of a motivating leader was a significant factor for commitment. For example, Derek’s adherence to the group’s regimen was partly attributed to the consistent encouragement he received from group leaders, who kept motivation high.
- **C4 - C5: Improvements in physical health motivate ongoing participation and commitment.**
  Physical gains acted as a feedback loop for motivation. Participants like Felicity noted that seeing measurable improvements in strength and endurance motivated them to continue attending and pushing themselves.
- **C4 - C6: Flexibility in group activities supports motivation, accommodating diverse needs and schedules.**
  Allowing flexibility in participation, such as offering alternative exercises, made it easier for members to stay committed despite personal limitations or fluctuating schedules, thereby maintaining motivation.
- **C4 - C8: Motivation is essential for long-term behavioural change, sustaining healthy habits.**
  Long-term changes in behaviour, such as maintaining a regular training schedule, were underpinned by sustained motivation. Participants cited motivation as the key factor that transitioned short-term engagement into a habitual commitment.
- **C4 - C9: Personal identity and self-perception affect motivation levels and sustained engagement.**
  How individuals saw themselves within the group influenced their commitment. Quinn's perception of himself as a dedicated member motivated him to attend consistently, reinforcing his commitment to the group's goals.
- **C4 - C10: Motivation is key to overcoming barriers and maintaining engagement.**
  High levels of motivation helped participants navigate challenges. For example, Kimberly’s determination to stay engaged, despite encountering health setbacks, demonstrated the role of motivation in overcoming barriers.

**C5: Physical Health and Capability**

- **C5 - C2: Physical health improvements contribute to better mental well-being.**
  Participants frequently mentioned that physical gains, such as increased strength and endurance, had a significant impact on their mental health. For example, Derek reported feeling less anxious and more positive as he noticed improvements in his physical capabilities, highlighting how physical progress supports emotional well-being.
- **C5 - C4: Health gains drive motivation and reinforce commitment to physical activity.**
  Physical improvements acted as a motivational feedback loop, reinforcing ongoing participation. Felicity highlighted how feeling physically stronger made her more motivated to attend sessions regularly, creating a virtuous cycle of health and engagement.
- **C5 - C7: Developing physical skills enhances overall capability and group engagement.**
  Skill acquisition was a key part of participants' physical development. Quinn’s progression from basic movements to more complex kettlebell exercises increased his confidence and encouraged more active engagement during sessions, showing that physical skills are foundational to deeper involvement.
- **C5 - C8: Physical health improvements support behavioural changes and maintenance.**
  As members became physically fitter, they were more likely to sustain new behaviours like regular training. Kimberly, for instance, transitioned to a healthier lifestyle that included more frequent and varied physical activities outside the group, suggesting that physical health gains underpin longer-term behaviour maintenance.
- **C5 - C10: Addressing physical challenges is essential for maintaining participation in activities.**
  Managing physical limitations or injuries was crucial for consistent engagement. Derek, who dealt with persistent back pain, often found ways to modify exercises to accommodate his physical condition, emphasising the importance of adaptive physical health strategies in overcoming participation barriers.

**C6: Practical Considerations and Flexibility**

- **C6 - C1: Practical flexibility aids in maintaining group cohesion.**
  The group's ability to adapt to members’ needs, such as flexible session times or changing training locations, was essential for maintaining cohesion. This adaptability allowed members like Megan to feel included and accommodated, reinforcing the group’s sense of unity.
- **C6 - C2: Practical elements reduce stress, supporting mental well-being.**
  Reducing logistical pressures through flexible arrangements lowered stress levels for participants. Derek appreciated the group’s leniency with attendance, as it allowed him to manage personal obligations without feeling guilty, contributing positively to his mental health.
- **C6 - C3: Effective leadership helps manage practical considerations within group settings.**
  Strong leadership was necessary to navigate practical challenges, such as ensuring access to training facilities or handling equipment. Sophia’s proactive approach in finding alternative training spots during rainy weather exemplified how leadership manages logistical concerns, ensuring group continuity.
- **C6 - C4: Flexibility supports motivation, accommodating diverse needs and schedules.**
  Adaptable training sessions allowed for varying levels of commitment, which helped maintain motivation. Kimberly noted that the group’s willingness to accommodate her schedule was a significant factor in keeping her engaged, showing that flexibility aids sustained motivation.
- **C6 - C9: Adaptable group activities contribute to a positive self-perception.**
  Allowing members to adjust their participation according to their comfort and ability levels contributed to a positive self-image. Quinn’s confidence grew as he saw that he could engage on his terms, reinforcing a sense of autonomy and control over his training journey.
- **C6 - C10: Practical barriers can be overcome with flexibility in planning and group structure.**
  Practical challenges, such as scheduling conflicts or access to facilities, were effectively mitigated through adaptive planning. For instance, the group’s ability to reschedule around participants’ commitments helped maintain high engagement, demonstrating the importance of logistical flexibility in overcoming barriers.

**C7: Skill Development and Knowledge Sharing**

- **C7 - C1: Social interactions encourage the sharing of skills and knowledge.**
  The social nature of the group fostered a culture of collective learning. Kimberly’s frequent tips and advice to other members about proper kettlebell form exemplify how social cohesion promotes knowledge exchange, leading to a more informed and capable group.
- **C7 - C2: Learning new skills boosts mental well-being and self-efficacy.**
  Participants often mentioned feeling more capable and self-assured as they acquired new skills. Felicity’s pride in mastering new exercises directly enhanced her confidence and reduced anxiety, demonstrating the connection between skill development and emotional health.
- **C7 - C3: Leadership fosters a culture of skill development and knowledge sharing.**
  Effective leaders like Sophia facilitated skill development by creating opportunities for knowledge exchange, such as organising mini-sessions focused on specific techniques. This leadership-driven focus on skill-building helped deepen the group’s collective expertise.
- **C7 - C5: Physical skill acquisition supports overall capability.**
  Learning new exercises and refining techniques improved members’ overall physical abilities. Derek’s progression in kettlebell skills enabled him to engage more fully in sessions, showcasing how physical competence boosts participation.
- **C7 - C8: Acquiring skills aids in long-term behaviour maintenance.**
  The development of practical skills was instrumental in sustaining new behaviours. Participants like Quinn highlighted that their newfound abilities made continued participation easier, showing that skills learned during the training acted as anchors for behavioural maintenance.
- **C7 - C9: Learning impacts identity, enhancing self-confidence and personal growth.**
  Acquiring and mastering new skills positively affected members’ self-perception. Megan expressed how learning complex movements made her feel more capable and competent, fostering a sense of personal achievement and growth within the group.
- **C7 - C10: Skill gaps can be barriers, but targeted training can overcome them.**
  Identifying and addressing skill gaps was crucial for sustained engagement. When members struggled with certain exercises, targeted guidance and demonstrations helped them overcome these barriers, ensuring that all participants could keep up with the group.

**C8: Behavioural Change and Maintenance**

- **C8 - C4: Sustained motivation is crucial for lasting behavioural change.**
  Motivation was identified as a key factor in sustaining new behaviours. Sophia’s consistent attendance and effort, driven by her sense of accountability to the group, underscored the importance of motivation in maintaining a long-term exercise habit.
- **C8 - C5: Health improvements reinforce new behaviours, making them sustainable.**
  As physical health improved, participants found it easier to maintain new habits. Felicity’s adherence to training increased as she noticed reductions in pain and improved mobility, demonstrating how physical progress can anchor behavioural change.
- **C8 - C7: Skill development supports consistent behaviour change.**
  Acquiring new skills was fundamental to maintaining behaviours over time. Megan’s mastery of specific exercises enabled her to integrate kettlebell training into her routine confidently, underscoring how skill proficiency can drive long-term adherence.
- **C8 - C9: Behavioural changes impact self-perception, reinforcing commitment.**
  Participants’ self-perception evolved as they sustained new behaviours, leading to a cycle of reinforcement. Derek saw himself as a committed member, which increased his drive to continue training, demonstrating how consistent behaviour influences identity.
- **C8 - C10: Addressing obstacles is necessary for maintaining behavioural changes.**
  Tackling challenges head-on was crucial for long-term adherence. Quinn’s strategy of modifying exercises to suit his physical limitations ensured that he could continue participating, showing that adaptation is key to maintaining behavioural shifts.

**C9: Identity and Self-Perception**

- **C9 - C1: A strong group identity positively influences individual self-perception.**
  Being part of a cohesive group positively shaped individual identities. Kimberly mentioned how her role in the group made her feel more valuable, reinforcing a sense of belonging and personal worth, which in turn encouraged further engagement.
- **C9 - C2: Improved emotional health enhances self-perception and identity.**
  As participants experienced emotional gains, their self-perception shifted positively. Megan noted that her anxiety decreased as she felt more capable, which led to a more confident and positive self-image over time.
- **C9 - C3: Leadership plays a pivotal role in shaping individual identities within the group.**
  Effective leadership contributed to a stronger sense of self among members. Sophia’s inclusive leadership style allowed quieter members like Quinn to find their voice and identity within the group, showing how leadership influences self-perception.
- **C9 - C4: Motivation is influenced by how individuals perceive themselves.**
  Participants’ commitment levels were directly tied to their self-perception. Quinn’s self-identification as a dedicated group member motivated him to attend regularly, suggesting that self-image is a driver for sustained engagement.
- **C9 - C6: Practical adaptations can shape how individuals see themselves within the group.**
  Flexibility in participation positively influenced self-perception. Felicity mentioned how adapting exercises to her comfort level made her feel more in control and capable, contributing to a more positive self-image.
- **C9 - C8: Behavioural changes influence identity, leading to a cycle of self-reinforcement.**
  Consistent behaviour over time had a transformative impact on self-identity. Megan, who initially doubted her physical abilities, grew to see herself as an active and fit individual through her consistent participation, reinforcing her commitment.
- **C9 - C10: Addressing identity challenges can overcome participation barriers.**
  Overcoming identity-based obstacles was key for long-term engagement. Quinn, who initially struggled with his perception as the ‘least fit’ member, overcame this challenge through positive reinforcement from the group, demonstrating the importance of tackling identity-related barriers.

**C10: Challenges and Barriers**

- **C10 - C1: Group cohesion aids in overcoming barriers.**
  The support network provided by a cohesive group helped participants navigate obstacles. For instance, when Megan faced scheduling conflicts, the group's understanding and flexibility allowed her to remain engaged despite logistical difficulties.
- **C10 - C2: Mental resilience is key to navigating challenges.**
  Emotional strength was essential for managing setbacks. Derek’s ability to handle personal challenges without withdrawing from the group was rooted in his increased mental resilience, which was bolstered by the supportive environment.
- **C10 - C3: Leadership can mitigate challenges and provide solutions.**
  Effective leadership played a crucial role in resolving group issues. Sophia’s approach to handling interpersonal tensions by fostering open communication showed how leadership can pre-empt and address challenges, maintaining group stability.
- **C10 - C4: High motivation levels are crucial for overcoming obstacles.**
  Strong motivation helped participants face barriers. Kimberly’s determination to keep training, even when health issues emerged, was a testament to the role of motivation in surmounting difficulties.
- **C10 - C5: Addressing physical limitations is vital for maintaining participation.**
  Adapting exercises to individual needs helped overcome physical challenges. Derek’s approach to modifying routines for his back issues allowed him to continue participating fully, highlighting the importance of accommodating physical barriers.
- **C10 - C6: Practical flexibility helps in mitigating logistical challenges.**
  Adjusting schedules and training environments addressed many logistical barriers, enabling continued participation. Felicity’s relief at the group’s adaptability during her recovery illustrated how flexibility can ease participation burdens.
- **C10 - C7: Skill development addresses and reduces barriers to participation.**
  Bridging skill gaps was crucial for group integration. When Quinn struggled with complex exercises, targeted support from other members improved his technique, ensuring he could continue participating without feeling out of place.
- **C10 - C9: Shifts in identity help individuals overcome participation challenges.**
  Changes in self-perception were instrumental in overcoming obstacles. Megan’s evolution from a hesitant participant to a confident group member allowed her to tackle challenges more effectively, illustrating how identity shifts impact engagement.
