## Supplementary File 6 for "Motivation, community, and resilience in older adults’ exercise: a three-year qualitative follow-up to the BELL trial" (from another version of this preprint: version 1, 2024-12-16)

##### Supplementary File 6: One-mode network for identifying connections among categories.

|  | C1 | C2 | C3 | C4 | C5 | C6 | C7 | C8 | C9 | C10 |
| --- | --- | --- | --- | --- | --- | --- | --- | --- | --- | --- |
| C1 | 1 | 1 | 1 | 1 | 0 | 1 | 1 | 0 | 1 | 1 |
| C2 | 1 | 1 | 0 | 1 | 1 | 1 | 1 | 0 | 1 | 1 |
| C3 | 1 | 0 | 1 | 1 | 0 | 1 | 1 | 0 | 1 | 1 |
| C4 | 1 | 1 | 1 | 1 | 1 | 1 | 0 | 1 | 1 | 1 |
| C5 | 0 | 1 | 0 | 1 | 1 | 0 | 1 | 1 | 0 | 1 |
| C6 | 1 | 1 | 1 | 1 | 0 | 1 | 0 | 0 | 1 | 1 |
| C7 | 1 | 1 | 1 | 0 | 1 | 0 | 1 | 1 | 1 | 1 |
| C8 | 0 | 0 | 0 | 1 | 1 | 0 | 1 | 1 | 1 | 1 |
| C9 | 1 | 1 | 1 | 1 | 0 | 1 | 1 | 1 | 1 | 0 |
| C10 | 1 | 1 | 1 | 1 | 1 | 1 | 1 | 1 | 0 | 1 |
