## Supplementary File 5 for "Motivation, community, and resilience in older adults’ exercise: a three-year qualitative follow-up to the BELL trial" (from another version of this preprint: version 1, 2024-12-16)

##### Supplementary File 5: Data representing the top 5 most prominent COM-B + intervention function combinations

### a) Education (*Psychological Capability*)

| **Code** | **Text Segment** | **Description** | **Theme** | **Intervention** |
| --- | --- | --- | --- | --- |
| CD8 | "Yeah, I did have depression, or medication for depression, it’s not only for depression, but it's also because I have a tendency to blow up like a volcano... I think was more negative, now it's not a negative." | Derek shares how his mental health, particularly his outlook on life, has improved since resuming physical activity, highlighting the psychological benefits of sustained engagement. | *Mental Health Improvement*: Highlights how physical activity has positively influenced mental health. | Offer education on mental health benefits of physical activity as part of a holistic approach to wellness. |
| CK22 | "We were in Oakey RSL... I said I’ll show you, and my daughter said [noises], and I’m down doing press-ups on the floor in the RSL club." | Kimberly provides a light-hearted anecdote showcasing her confidence and strength, doing push-ups in a public setting as a reflection of her ongoing physical prowess. | *Confidence and Strength*: Demonstrates the confidence and pride gained from physical achievements. | Use real-life examples of participants’ strength improvements to motivate others. |
| CS32 | "I think there could be so much more. It’s just a mentality." | Sophia reflects on the limitations older individuals place on themselves, seeing potential for much more physical activity. | *Mental Barriers*: Sophia sees older people limiting their own potential for physical activity. | Educate members on how consistent training contributes to long-term resilience. |

### b) Enablement (*Social Opportunity*)

| **Code** | **Text Segment** | **Description** | **Theme** | **Intervention** |
| --- | --- | --- | --- | --- |
| CD14 | "If Kimberly’s doing it, Julia’s doing it, join along; that’s the way it [always] was at the gym." | The group’s collective approach to training creates a supportive dynamic, where individuals follow each other’s lead, maintaining participation through mutual encouragement. | *Collective Engagement*: Highlights the follow-the-leader dynamic in fostering group participation. | Encourage experienced members to serve as informal leaders, guiding newcomers and fostering collective engagement. |
| CF2 | "I'm a social person, I love meeting people, talking, chatting, having a coffee and that's why I just go to the coffee." | Felicity expresses that her primary motivation for attending the group is social interaction rather than exercise, highlighting the importance of social connections. | *Social Engagement*: Emphasises social interaction as a primary motivation for participation. | Structure social gatherings around exercise sessions to build a supportive community that encourages regular attendance. |
| CJ25 | "I don't mind [goblet] squats. Quinn hates squats. If Sophia is there, normally she's the boss." | Julia mentions specific exercises she enjoys, highlighting the influence of Sophia's leadership in maintaining structure during the group sessions. | *Physical and Social Accountability*: Describes how the group serves as an accountability mechanism for both physical activity and social engagement. | Encourage members to take turns leading activities, promoting respect for each other's exercise preferences. |

### c) Environmental Restructuring (*Social Opportunity*)

| **Code** | **Text Segment** | **Description** | **Theme** | **Intervention** |
| --- | --- | --- | --- | --- |
| CD2 | "The training’s great but socially it's a different sort of thing; you get to know the person." | Derek notes the dual benefit of the group sessions, where both training and socialising contribute to a sense of well-being and connection. | *Social Connection*: Highlights both physical training and social interaction as sources of well-being. | Encourage opportunities for socialising within training groups to enhance connection and well-being. |
| CF19 | "To me, it's important to then go and have a coffee together or something. The ones that I keep doing, like meeting up with the Wednesday friends group is the social coffee." | Felicity emphasises the importance of the social aspect of group activities, particularly the coffee meetups, which are central to her continued engagement, reinforcing the significance of social opportunity. | *Importance of Socialising*: Emphasises that the social aspect of the group is key to her continued engagement. | Schedule regular social gatherings, like coffee meetups, after exercise to enhance social interaction and long-term engagement. |
| CM42 | "There's no judgement, they don’t say like ‘why don’t you come to kettlebells?’ it’s ‘oh, you’re here!’" | Megan appreciates the non-judgmental attitude within the group, where members are welcomed regardless of participation in the kettlebell exercises. | *Non-Judgmental Environment*: Appreciates the group’s welcoming, non-judgmental attitude. | Foster a non-judgemental group culture for inclusivity. |

### d) Training (*Physical Capability*)

| **Code** | **Text Segment** | **Description** | **Theme** | **Intervention** |
| --- | --- | --- | --- | --- |
| CF11 | "The fact that I can do it, I can do it, I can push myself and do more than I thought I could be bothered doing it." | Felicity expresses a newfound confidence in her physical capability, underscoring the impact of the trial on her self-efficacy and motivation to engage in exercise. | *Self-Efficacy*: Highlights a newfound confidence in physical abilities. | Use feedback and progressive exercises to reinforce physical competence and confidence. |
| CJ4 | "It just made it easy, you could feel how strong you were, and even in opening jars..." | Julia values the tangible improvements in physical strength, noting how the trial improved her ability to perform everyday tasks, reinforcing the impact of kettlebell training on physical functioning. | *Functional Strength*: Focuses on the ease of performing everyday activities, such as opening jars, thanks to increased strength. | Include exercises that strengthen physical tasks (like grip), which are commonly needed daily. |
| CS14 | "The improvement in strength when the chips are down and things go wrong can come back quickly if you keep at it." | Sophia highlights the resilience built through kettlebell training and its role in helping her maintain strength despite health setbacks. | *Resilience through Training*: Training helps maintain strength and resilience despite health setbacks. | Regularly revisit exercise fundamentals to build participant confidence in their skills. |

### e) Education (Reflexive Motivation)

| **Code** | **Text Segment** | **Description** | **Theme** | **Intervention** |
| --- | --- | --- | --- | --- |
| CD19 | "We did learn how to use kettlebells as I said before, which was good. I think that just gave me the motivation to carry on." | Derek emphasises how acquiring the proper technique during the BELL trial provided him with the confidence and motivation to continue training. | CD19 – Confidence Building: Reflects on how learning proper kettlebell techniques sustained motivation. | Share personal success stories or motivational moments from the training to help sustain interest and dedication to the program. |
| CK13 | "The kettlebells 2 instigated it. Without the kettlebells, I'd probably still be..." | Kimberly highlights the significant role kettlebell training played in restoring her mobility, even though her challenges predated the trial. | CK13 – Motivation and Mobility: Emphasises kettlebell training’s role in restoring mobility and drive. | Educate participants on the benefits of kettlebell training for mobility restoration, reinforcing its role in continued engagement. |
| CK39 | "If I hadn't signed up for the training it wouldn't have happened." | Kimberly attributes her significant lifestyle changes directly to the kettlebell training and acknowledges the importance of that initial decision to join the trial. | CK39 – Importance of Initial Participation: Acknowledges the significance of joining the trial for subsequent lifestyle changes. | Highlight the importance of initial commitment to the program as a catalyst for sustained lifestyle change. |
