## Supplementary File 4 for "Motivation, community, and resilience in older adults’ exercise: a three-year qualitative follow-up to the BELL trial" (from another version of this preprint: version 1, 2024-12-16)

##### Supplementary File 4. Clustering codes into categories

|  |  | **Codes** |
| --- | --- | --- |
| Categories | C1 – Social Cohesion and Support | CD1, CD2, CD10, CD12, CD18, CD20, CF2, CF4, CF6, CF7, CF8, CF12, CF13, CF19, CF25, CF47, CF48, CF49, CJ1, CJ5, CJ8, CJ9, CJ16, CJ18, CJ22, CJ29, CJ30, CK2, CK4, CK6, CK10, CK12, CK15, CK21, CK28, CK42, CK44, CM1, CM9, CM14, CM15, CM42, CM47, CM48, CQ1, CQ5, CQ6, CQ7, CQ9, CQ10, CQ20, CQ21, CQ23, CQ33, CQ42, CQ51, CQ62, CS1, CS2, CS7, CS18, CS21, CS22, CS27, CS30, CS42, CS45, CS50, CS59, CS61, CS63 |
|  | C2 – Mental and Emotional Well-being | CD6, CD11, CF14, CF27, CJ2, CJ10, CJ40, CJ42, CK8, CK14, CK16, CK40, CM7, CM8, CM52, CQ3, CQ4, CQ12, CQ15, CQ31, CQ46, CS6, CS13, CS15, CS40, CS57, CS66 |
|  | C3 – Leadership and Group Dynamics | CD14, CD15, CD24, CD28, CD29, CF5, CF26, CF28, CF39, CF40, CF41, CJ7, CJ24, CJ25, CJ32, CJ48, CK5, CK13, CK20, CK22, CK26, CM2, CM11, CM12, CM13, CM24, CM37, CM43, CM44, CM45, CQ8, CQ13, CQ22, CQ30, CQ34, CQ37, CQ48, CQ57, CS3, CS10, CS23, CS25, CS37, CS46, CS64, CS67, CS70 |
|  | C4 – Motivation and Commitment | CD5, CD13, CD19, CD25, CD26, CF1, CF11, CF15, CF24, CF35, CF36, CF50, CF5, CJ6, CJ15, CJ19, CJ34, CJ43, CK3, CK9, CK19, CK29, CK41, CM4, CM5, CM6, CM19, CM35, CM50, CQ2, CQ14, CQ18, CQ19, CQ25, CQ35, CQ38, CQ39, CQ44, CS5, CS16, CS19, CS29, CS32, CS34, CS36, CS47, CS49, CS51, CS58, CS62 |
|  | C5 – Physical Health and Capability | CD3, CD4, CD7, CD9, CF9, CF10, CF20, CF21, CF29, CF30, CF33, CJ3, CJ4, CJ11, CJ12, CJ41, CK1, CK7, CK23, CK27, CK34, CK47, CM18, CM20, CM22, CM26, CQ11, CQ17, CQ27, CQ28, CQ29, CQ49, CQ53, CQ64, CS4, CS26, CS28, CS35, CS41, CS53, CS56, CS68, CS71 |
|  | C6 – Practical Considerations and Flexibility | CD21, CD22, CD23, CD27, CF31, CF34, CJ14, CJ20, CJ28, CJ33, CJ47, CK11, CK18, CK24, CK30, CK31, CK35, CK37, CK39, CK43, CM3, CM21, CM23, CM51, CQ16, CQ24, CQ26, CQ40, CQ50, CQ55, CS20, CS38, CS43, CS44, CS65 |
|  | C7 – Skill Development and Knowledge Sharing | CD16, CD17, CF42, CF43, CJ13, CJ23, CJ45, CK17, CK36, CM27, CM28, CM29, CM30, CM31, CQ32, CQ41, CQ43, CQ54, CQ56, CQ58, CS17, CS52, CS69 |
|  | C8 – Behavioural Change and Maintenance | CD8, CF3, CF22, CF37, CF44, CJ17, CJ26, CJ39, CJ44, CK25, CK33, CK38, CK45, CM32, CM34, CM39, CM41, CM40, CQ36, CQ45, CQ59, CQ60, CS8, CS14, CS33, CS39, CS47, CS60 |
|  | C9 – Identity and Self-Perception | CF32, CF45, CJ21, CJ35, CJ46, CK46, CM16, CM17, CM25, CS9, CS32, CS48, CS55 |
|  | C10 – Challenges and Barriers | CF16, CF17, CF18, CF23, CF38, CF46, CJ27, CJ31, CJ36, CJ37, CJ38, CJ49, CK32, CM10, CM33, CM36, CM38, CM49, CQ61, CS11, CS12, CS24, CS31, CS54 |
