## Supplementary File 3 for "Motivation, community, and resilience in older adults’ exercise: a three-year qualitative follow-up to the BELL trial" (from another version of this preprint: version 1, 2024-12-16)

##### Supplementary File 3. Interview guide (b)

- What did you **enjoy** the most from the trial, and what would you say was the biggest **benefit** for you in completed it?
- Do you feel you have **maintained** the benefits from the trial?
- Tell me about the ‘Wednesday friends’ group – how it started, how you came to be a part of it, and what you think has kept it going all this time.
- Can you tell me about the **friendships** you’ve made in the group, and what that’s meant to you.
- How important have the **social connections** been to you?
- Is there anything else about that group that’s been particularly **important** to you?
- What has kept you **motivated** to continue training all this time?
- Can you describe the **group dynamics** during the training sessions? How do these dynamics influence your engagement in the activity?
- Would you still be training if you weren’t a part of the group?
- How do you feel on days when you can't attend the kettlebell training session? Does this influence your motivation to attend the next session?
- In what ways, if any, have the training sessions influenced your relationships with **family and friends** outside the group?
- What **challenges**, if any, have you faced in maintaining your commitment to the kettlebell training sessions? How have you and the group addressed these challenges
- Have you experienced any changes in your overall **health and well-being** since you began participating in the kettlebell training sessions? If so, can you describe these changes?
- How do you perceive the impact of the kettlebell training sessions on your **physical, mental, and emotional well-being**?
- Has your perspective on **physical activity** and **healthy ageing** changed since you began the kettlebell training? Could you explain how?
- Could you share a **memorable** experience during these training sessions that profoundly affected you or your commitment to the activity?
- How has your participation in the kettlebell training sessions influenced your **self-confidence**, if at all?
- What **advice** would you give to other older adults who might be considering participating in a similar group-based physical activity program?
- What do you tell your friends and family about the group?
- Tell me about your confidence, psychologically, using kettlebells - are you able to give me an example?
- How did what you learnt from the trial set you up to do what you’ve been doing as part of your weekly sessions?
- How did you feel the trial **physically prepared** you for training in the park every week without any supervision and guidance?
- Can you talk to me about the **knowledge and skills** you got from the trial, that you feel has helped you continue training as part of the group?
- How has your training with the kettlebells changed since the trial finished; are you using them differently?
- What **factors** would you say have supported and helped you to participate in the weekly training sessions? e.g., it’s convenient to get to, right time of day, you’re being encouraged to do it at home etc.
- What sorts of things have **got in the way** of you training, or stopped you from being able to do it altogether?
- For the benefit of the people who will be reading about what you’ve been doing as part of the group training, can you tell me a little about **where** the group trains every week, how easy or difficult it has been to get everyone **together**, and what’s worked well to keep it running for the last three years?
- What sort of **support** have you had to continue training and be a part of the Wednesday Friends Group?
