## Supplementary File 2 for "Motivation, community, and resilience in older adults’ exercise: a three-year qualitative follow-up to the BELL trial" (from another version of this preprint: version 1, 2024-12-16)

##### Supplementary File 2. Interview guide (a)

| **Objective** | **Interview questions** | **Anticipated outcomes** |
| --- | --- | --- |
| **Motivation**  To investigate the motivation behind the continued participation of these individuals in kettlebell training sessions. | Tell me what you remember about your experience in the BELL trial, and what motivated you to take part in it. | Identification of factors that contribute to the sustained engagement of the small group of former BELL trial participants in their independent group-based kettlebell training sessions. |
|  | What did you enjoy the most from the trial, and what would you say was the biggest benefit for you in having completed it? [benefits] |  |
|  | Tell me about the ‘Wednesday friends’ group – how it started, how you came to be a part of it, and what you think has kept it going all this time. |  |
|  | Can you tell me about the friendships you’ve made in the group, and what that’s meant to you [social connections] How important have the social connections been to you? Is there anything else about that group that’s been particularly important to you?  What has kept you motivate to continue training all this time?  *explore this more deeply in relation to friendships, meaningful activity, lifestyle changes, personal confidence…* |  |
| To identify the role of group dynamics and social support in fostering long-term engagement in the physical activity. | Can you describe the group dynamics during the training sessions? How do these dynamics influence your engagement in the activity? | Better understanding of the role of social connections, group dynamics, and perceived health benefits in promoting long-term participation in group-based physical activities among older adults. |
|  | *ask about what happens in the group – is there always a clear leader, does the format stay the same, does everyone do the same thing at the same time* |  |
|  | Would you still be training if you weren’t a part of the group? |  |
|  | *also want to know are there better groups /days than others* |  |
|  | How do you feel on days when you can't attend the kettlebell training session? Does this influence your motivation to attend the next session? |  |
|  | In what ways, if any, have the training sessions influenced your relationships with family and friends outside the group? |  |
|  | *May consider asking about any other group programs they attend – are they similar/ different?* |  |
|  | What challenges, if any, have you faced in maintaining your commitment to the kettlebell training sessions? How have you and the group addressed these challenges |  |
|  | Have you experienced any changes in your overall health and well-being since you began participating in the kettlebell training sessions? If so, can you describe these changes? |  |
|  | How do you perceive the impact of the kettlebell training sessions on your physical, mental, and emotional well-being? |  |
|  | *Also want to know whether the benefits from the original trial are being maintained* |  |
|  | Has your perspective on physical activity and healthy ageing changed since you began the kettlebell training? Could you explain how? |  |
|  | Could you share a memorable experience during these training sessions that profoundly affected you or your commitment to the activity? |  |
|  | How has your participation in the kettlebell training sessions influenced your self-confidence, if at all? |  |
|  | What advice would you give to other older adults who might be considering participating in a similar group-based physical activity program? |  |
|  | *May want to ask, what do you tell your friends and family about the group?* |  |
| **Capability** | Tell me about your confidence, psychologically, using kettlebells - are you able to give me an example? [psychological capability] | |
|  | How did what you learnt from the trial, set you up to do what you’ve been doing as part of your weekly session? [knowledge] | |
|  | How did the trial physically prepare you for training in the park every week without any supervision and guidance? [physical capability] | |
|  | Can you talk to me about the knowledge and skills you got from the trial, that you feel has helped you continue training as part of the group? [knowledge and skills] | |
|  | How has your training with the kettlebells changed since the trial finished; are you using them differently? [improved capabilities through training] | |
| **Opportunity** | What factors would you say have supported and helped you to participate in the weekly training sessions? | |
|  | *if a prompt is needed*… for example – it’s convenient to get to, right time of day, you’re being encouraged to do it at home – anything you can thing of that’s made it easy to do [facilitating external factors] | |
|  | What sorts of things have got in the way of you training, or stopped you from being able to do it altogether? [hindering external factors] | |
|  | For the benefit of the people who will be reading about what you’ve been doing as part of the group training, can you tell me a little about where the group trains every week, how easy or difficult it has been to get everyone together, and what’s worked well to keep it running for the last three years? [availability] | |
|  | What sort of support have you had to continue training and be a part of the Wednesday friends’ group? [environmental or social influences] | |
|  | What factors would you say have supported and helped you to participate in the weekly training sessions? | |
