## Supplementary File 1 for "Motivation, community, and resilience in older adults’ exercise: a three-year qualitative follow-up to the BELL trial" (from another version of this preprint: version 1, 2024-12-16)

### Supplementary Information

##### Supplementary File 1. BQQRG guidelines for qualitative research

| **Article section of element** | **Alignment with BQQRG guidelines** |
| --- | --- |
| Writing style | This manuscript utilises humanised and subjective language to convey the social and psychological dynamics within the Wednesday Friends group. The writing incorporates longitudinal insights drawn from ongoing interaction with participants’ shared experiences. Vivid, contextualised descriptions replace detached reporting, framing the study’s findings in a way that foregrounds the social context and lived experience of the group. |
| Terminology | Key terms, such as *intrinsic motivation*, *self-determined motivation*, and *reflexive engagement*, are clearly defined to ensure clarity and alignment with participants' experiences in sustaining exercise. Reflexive engagement, acknowledging the lead researcher’s ongoing relationship with the group, is integrated into the methods to illustrate nuanced understanding and interpretation. Terminology choices are informed by qualitative research standards and adapted to the context of older adult exercise motivation, avoiding stigmatising language and explaining all contested terms as recommended. |
| Title | The title was revised to align with the BQQRG guidelines, capturing the main topic, key themes, and timeframe of the study. This title reflects the core storyline—sustained motivation, community bonds, and resilience among older adults in a group exercise context. Keywords were included in the first part for discoverability and relevance to potential readers |
| Abstract and keywords | The abstract is structured to convey the essence of the study, summarising context, methodology, and main findings in an engaging format. Key qualitative frameworks, including the COM-B model and Four Cs model, are highlighted to reflect the study's approach to understanding older adults' engagement in group exercise. Keywords extend beyond the title to include theoretical and methodological terms that improve discoverability and signal the study’s qualitative focus. |
| **Introduction** | |
| Background and rationale | The Introduction contextualises the study within the broader discourse on exercise adherence for older adults, engaging with prior research from a perspective that values participants' subjective experiences. Existing quantitative findings are discussed as part of a partial narrative, with this study positioned as contributing a qualitative perspective. Explicit connections to ongoing societal conversations around aging and health further ground the rationale for this research, adhering to BQQRG guidance on robust contextualisation and theoretical framing. |
| Research question, aims, and/or purpose | The aims are explicitly stated and aligned with the qualitative, experiential focus of the study. The manuscript clearly situates this work as a qualitative follow-up to the BELL trial, with a unique focus on exploring underlying motivations and long-term engagement in self-sustained group-based exercise. |
| ‘Owning your perspectives’ – discussion of theory | The methodology includes a description of Critical Realism as the guiding meta-theoretical framework, balancing subjective insights with inferred influences. COM-B and thematic analysis are utilised within this framework to explore both visible behaviours and underlying dynamics, ensuring congruence between theoretical stance and analytic approach. This reflexive framework acknowledges that participants’ experiences, while interpreted by the researcher, reveal layered factors sustaining engagement in group exercise. |
| ‘Owning your perspectives’ – discussion of reflexivity | The methodology includes a clear statement on reflexivity, acknowledging NM’s extended engagement with the participant group from the original BELL trial through ongoing interactions. This connection provided depth of insight but required continuous reflexive practice to maintain an interpretive balance between familiarity and objectivity. NM’s reflexivity is discussed in terms of recognising potential biases and consciously examining assumptions throughout data collection and analysis, demonstrating alignment with BQQRG's emphasis on critical reflexivity, researcher positioning, and interpretive awareness. |
| **Methodology** |  |
| Research design/approach/methodology | The Methods section clearly articulates the use of a Critical Realist framework and reflexive thematic analysis (RTA), aligning with the study’s focus on exploring both observable behaviours and underlying mechanisms. Reflexivity is integrated throughout, with NM’s prolonged engagement ensuring balanced interpretation, while the COM-B model and the Four C’s model provide a pluralistic approach that addresses both individual and social dimensions of participant experiences, adhering to BQQRG’s emphasis on methodological congruence and transparency. |
| Participants/data sources/empirical materials | This study clearly describes the recruitment and inclusion criteria for the Wednesday Friends group members, ensuring transparency in participant selection. NM’s extended engagement with the group is acknowledged to underscore the reflexive interpretation of participant insights, enhancing the contextual understanding of sustained engagement without compromising participant anonymity. |
| Ethical approval and ethical code/principles followed | Ethical approval details, the informed consent process, and participant confidentiality measures are clearly stated. Researcher-participant dynamics are transparently addressed, acknowledging the influence of NM’s prior relationship with participants on study interpretation and reflexive practices. Additional accessibility considerations were not required given the nature of the study and participant group. |
| Data generation | The document provides a detailed overview of the data generation process, including selection and recruitment rationale, protocol development aligned with the COM-B model, and ethical considerations in participant engagement. Data preparation is clearly described, ensuring accuracy and respecting participant anonymity. |
| Data analysis procedures | The analysis followed an inductive approach, guided by Braun & Clarke’s (2021a) thematic analysis and the Four C’s model (Christodoulou, 2023). Data were coded in a flexible, open-ended manner, with codes developed from the data to explore participants' engagement mechanisms. Analysis was conducted individually by NM, who maintained reflexivity to mitigate potential biases. Supplementary files outline the clustering of codes, the identification of key themes, and the relationships between categories. |
| Quality practices | This study employs Big Q congruent quality practices by incorporating reflexivity throughout the analysis. NM, as the primary analyst, engaged in ongoing self-reflection to mitigate bias arising from prior relationships with participants. No postpositivist practices, such as triangulation or consensus coding, were used. Instead, the study’s quality is ensured through continuous engagement with the data, member reflections, and the use of supplementary materials to provide transparency in the analytic process. Limitations related to the dataset and participant group, including limited diversity of participants, are acknowledged, but these factors are considered in the context of the study's qualitative aims. |
| **Analysis** | |
| Structure, number, and names of patterns / themes / categories / discourses / narratives (etc.) | The analysis follows a clear structure informed by the Four C’s model. We identified ten unified categories, which provided a focused yet comprehensive exploration of the data. The category names were chosen to reflect the study’s key themes, ensuring both clarity and depth in the presentation of the analysis. The analytic structure is coherent, avoiding unnecessary fragmentation, and the number of codes is not presented as a measure of validity. |
| Overview of, or introduction to, analysis, if relevant | An analytic map and associated tables visually present the relationships between the ten unified categories, aligning with the subheadings used throughout the analysis section. The pattern names reflect the core themes of the study and are consistent with the structure previewed in the analysis, ensuring clarity and coherence in presenting the findings. |
| Theme/category/discourse/ narrative (etc.) conceptualisation, where relevant | The categories in this study were conceptualised in line with the Four C’s model (Christodoulou, 2023) and the principles of reflexive thematic analysis (Braun & Clarke, 2021a). Each category reflects a central organising concept that captures the shared meaning within the data. Any divergences in the conceptualisation of the categories, such as the integration of the COM-B model alongside the Four C’s model, were made to align with the study’s specific aims and ensure methodological consistency, ultimately enhancing the depth of the analysis. |
| Analytic depth | Utilising the Four C’s model, retroductive reasoning is employed to identify causal mechanisms that explain the connections between categories and participants' engagement. NM’s active and reflexive engagement with the data ensured the analysis was grounded in participant experiences, providing a deep understanding of the interrelated factors driving sustained group involvement. |
| Use of data extracts, where relevant | The study includes a sufficient range of data extracts that are well-contextualised and aligned with the analytic narrative. The extracts, drawn from a range of participants, illustrate the key themes identified in the analysis, such as Mental Health Improvement, Confidence and Strength, and Social Engagement. The balance between analytic narrative and data extracts is well-maintained, with the narrative driving the analysis and the extracts serving to illustrate key points. Anonymisation is appropriately handled, and the treatment of language aligns with the Critical Realist framework, reflecting the complexity of participant experiences. |
| Integration of analysis with existing research and theory | The study employs a range of data extracts that are appropriately contextualised to illustrate key themes. These extracts, drawn from various participants, are carefully linked to the analytic narrative, supporting and enriching the analysis. The balance between analytic narrative and data extracts is maintained, with the narrative guiding the interpretation and the extracts serving to illustrate key points. Anonymity of participants is strictly upheld, and the treatment of the data aligns with the Critical Realist framework, reflecting the complexity of participants' experiences. |
| Integration of analysis with existing research and theory | The study integrates relevant theories, including Self-Determination Theory, Resilience Theory, and Social Capital Theory, to explain participant behaviours and engagement in group-based exercise. It situates the findings within existing literature, deepening our understanding of how social support and intrinsic motivation contribute to long-term exercise adherence in older adults. |
| **Discussion/Conclusion** | |
| Analytic conclusions and contributions | The study provides clear conclusions linking social support, resilience, and intrinsic motivation to sustained engagement in exercise programs. It offers practical insights for healthcare providers and policymakers on structuring interventions, making a valuable contribution to the literature on exercise adherence among older adults. |
| Study evaluation/reflection | The study reflects on the strengths and limitations of the study, particularly the researcher’s influence, which enriched the analysis. The iterative nature of the analysis are acknowledged, ensuring that the findings are grounded in participants’ experiences. |
| Researcher reflexivity | Reflexivity is emphasised, acknowledging how the researcher’s background in community health interventions shaped the study. The manuscript highlights that researcher influence is inherent and beneficial in Big Q methodologies, adding depth to the analysis while maintaining focus on participant perspectives. |
| Implications for future research, policy and practice, where relevant | The findings suggest that social cohesion, intrinsic motivation, and resilience should be central to the design of future community-based health interventions for older adults. The study offers practical implications for public health policy and program design, with suggestions for future research to test these strategies in different contexts. |
